# Modelling of COVID-19 vaccination strategies and herd immunity, in scenarios of limited and full vaccine supply in NSW, Australia

**DOI:** 10.1101/2020.12.15.20248278

**Authors:** C Raina MacIntyre, Valentina Costantino, Mallory Trent

## Abstract

Several vaccines for SARS-CoV-2 are expected to be available in Australia in 2021. Initial supply is likely to be limited, and will require a judicious vaccination strategy until supply is unrestricted. If vaccines have efficacy as post-exposure prophylaxis (PEP) in contacts, this provides more policy options. We used a deterministic mathematical model of epidemic response with limited supply (age-targeted or ring vaccination) and mass vaccination for the State of New South Wales (NSW) in Australia. For targeted vaccination, the effectiveness of vaccinating health workers, young people and older adults was compared. For mass vaccination, we tested varying vaccine efficacy (VE) and distribution capacities. With a limited vaccine stockpile of 1 million doses in NSW, if there is efficacy as PEP, the most efficient way to control COVID-19 will be ring vaccination, however at least 90% of contacts per case needs to be traced and vaccinated. Health worker vaccination is required for health system resilience. Age based strategies with restricted doses make minimal impact on the epidemic, but vaccinating older people prevents more deaths. Herd immunity can only be achieved with mass vaccination. With 90% VE, herd immunity can be achieved by vaccinating 66% of the population. A vaccine with less than 70% VE cannot achieve herd immunity and will result in ongoing risk of outbreaks. For mass vaccination, distributing at least 60,000 doses per day is required to achieve control. Slower rates of vaccination will result in the population living with COVID-19 longer, and higher cases and deaths.

## Introduction

Coronavirus disease 19 (COVID-19) has caused an unprecedented pandemic, with catastrophic health, social and economic impacts ^1^. Without available drugs or vaccines, nations require ongoing or intermittent use of non-pharmaceutical approaches such as lockdowns, border closures and social distancing to control COVID-19^2,3^. In addition, case finding through increased testing and surveillance, with contact tracing and quarantine, are the mainstays of epidemic control ^4^. While Australia has achieved good control over the pandemic through border closures, testing, contact tracing and hotel quarantine, the risk of intermittent outbreaks of COVID-19 will continue until an effective vaccine is available ^3^. The societal impact of ongoing restrictions and border closure cannot be under-estimated.

There is a massive global effort to fast-track the development of COVID-19 vaccines ^5,6^. Currently, there are 48 COVID-19 vaccine candidates undergoing clinical evaluation, with 11 already in phase 3 clinical trials ^7^. Preliminary reports from phase 3 trials suggest that mRNA vaccines have over 90% efficacy against COVID-19 infection ^8,9^. The ChAdOx1 nCoV-19, the largest component of the planned Australian vaccine stockpile, has efficacy of 62% against symptomatic infection in the intended two-dose schedule ^10^. On December 11 it was announced that the second largest component of the Australian stockpile had been withdrawn from further development ^11^. A small proportion of the Australian vaccine plan includes the BNT162b2 mRNA vaccine, which has 95% efficacy against symptomatic infection ^11,12^. As more vaccine candidates become available through 2021, we will continue to see variation in efficacy and safety between them.

It is likely there will be initial vaccine shortages. Thus, an effective vaccination strategy will need to be developed to best utilise limited vaccine supply in the early phases of vaccine rollout ^13^, with a plan for expanded vaccination at a later stage.

Several COVID-19 vaccination strategies have been proposed, ^13^ such as prioritising healthcare and aged care workers and other frontline responders at high risk of disease transmission, and sociodemographic groups at significantly higher risk of severe disease, such as older adults or people with high risk chronic health conditions ^13,14^. However, prioritizing children or young people may impact transmission more, since vaccines are more effective in younger people, and transmission is highest in young adults^15^.

Alternatively, a ring vaccination strategy could be utilized, which involves identifying the close contacts of a confirmed case and vaccinating them. This strategy was used effectively against Ebola, and also smallpox in settings where mass vaccination was not possible, despite efficacy being half that of primary prevention ^16–19^. Many vaccines including measles, hepatitis A and smallpox, are effective as post-exposure prophylaxis (PEP) and can be given to contacts during an outbreak, albeit with lower efficacy than primary prevention ^20^. Whether COVID-19 vaccines will be effective as PEP is unknown as yet, but may well be given the long incubation period ^21^.

In Australia, the stated priority groups for early COVID-19 vaccination are: 1) individuals with increased risk of severe disease, such as older adults, Aboriginal and Torres Strait Islander people, and those with high risk chronic conditions; 2) individuals at higher risk of disease, such as health and aged care workers; and 3) individuals working in critical services ^22^. The aim of this paper was to model the impact of various COVID-19 vaccine strategies under a limited supply scenario, as well as varied vaccine efficacy and speed of vaccination for mass vaccination, on COVID-19 case numbers and mortality in New South Wales, the most populous state of Australia.

## Methods

A previously published age structured deterministic compartmental model for Covid-19 spread ^23^ was modified to test several possible vaccination strategies under limited or unlimited supply assumptions, as described in more detail below. We used the NSW population and age distribution from 2020 ^24^, stratified into 16 five-year age groups from 0 to 74 years old and the last age group comprising people 75 years and over.

The model moves the population in 16 mutually exclusive compartments, each divided by the 17 age groups: susceptible (S), latent, not infectious (E), pre-symptomatic infectious and diagnosed (E^t^), pre-symptomatic infectious and undiagnosed (E^u^), contacts traced (C) symptomatic, high infectiousness, diagnosed (I1) and undiagnosed (I2), asymptomatic, high infectiousness (A1), symptomatic, lower infectiousness, previous diagnosed (I11) and undiagnosed (I22), asymptomatic low infectiousness (A2), home isolated (Q), hospitalized (H), individuals admitted to intensive care (ICU), recovered (R), successfully vaccinated (V) and COVID-19 deceased people (D). The duration of stay in each compartment determines the rate at which people move from one compartment to another for the epidemiological disease states. The duration of the latent period is assumed to be 5.2 days ^25^, of which the last two days before symptoms onset are considered infectious ^26^.

The transmission was distributed over the infectious period with 44 % of transmissions occurring in the last two days of the pre-symptomatic state. We assume that the viral load is very high on the first day of symptoms and then decreases to a lower infectious level, spread over the following 6 days ^26^, so 36% of transmissions occur in the first day of symptoms and 20% in the following 6 days of symptoms ^26^. Of the total people infected, 35% are considered asymptomatic and never develop symptoms, however we assumed that asymptomatic cases are as equally infectious as symptomatic ones ^27–29^. In conjunction with vaccination, other disease control interventions such as case finding, contacts tracing, quarantine, isolation and hospital, ICU treatments were included, however we did not include additional measures such as universal masking or lockdowns, neither of which are in effect in NSW.

The force of infection that moves people from susceptible to infected is an age specific rate calculated as a combination of average age specific number of close contacts per day in Australia ^30^, the proportion of infectious people and the probability of infection per contact, which is estimated to reproduce an R0 of 2.5 ^28^. Once infected, a person enters the latent compartment, and after 3.2 days will become infectious and pre-symptomatic for 2 days, where if traced will be quarantined with a 50% reduction in transmissions ^31^ and upon symptoms onset will take 1 day on average to get isolated with no more transmissions after isolation. However, if an infected person is not traced in latency, it will take 5 days to get isolated following symptom onset ^32^. Hospitalizations and admission to ICU follow age-specific rates ^33^. The model assumes that 80% of all close contacts are traced and quarantined for 14 days and 90% of the symptomatic cases are isolated after 5 days.

There are no data on COVID-19 vaccines as PEP, but early data from phase three trials indicate efficacy in excess of 90% as primary prevention of mRNA vaccines ^8^. Vaccine efficacy was set at 90% in the base case scenario if a person is susceptible and 45% if the vaccine is given in the early latent epidemiological state as PEP. All parameters used in the model, as well as differential equations are described in the supplementary materials.

In order to test the most effective vaccination distribution for outbreak control, we used a hypothetical epidemic in NSW with 100 symptomatic people and 250 untraced latent infected people as the starting conditions, so that we could test the effectiveness of different vaccination strategies in an epidemic scenario.

### Vaccination strategies tested

1. *Limited vaccine supply* (1 million doses) given to targeted age groups
2. *Limited vaccine supply*, ring vaccination (contact tracing and vaccination of contacts)
3. *Unlimited supply and mass vaccination* assuming enough doses to vaccinate the entire NSW population

In the limited supply scenario, we assumed a limited stock of only 2 million doses (for 1 million people in a two-dose schedule) for NSW as an estimate of initial supply. For mass vaccination we assumed a delivery capacity of 50,000-300,000 doses per day ^34^. We explored 3 different scenarios with a limited vaccine supply:

1. Vaccine delivered in 8 days to 1 million young people (age group 10-29);
2. Vaccine delivered in 8 days (first dose) to 1 million older people (age group 65+);
3. Vaccinate 125,000 HCWs with first dose and over the following 7 days 875,000 people aged 10-29;

The two dose schedule was assumed to be given with 21-28 days between dose one and two. ^10,12^ The health care workers (HCWs) for NSW were estimated from the 2019 estimation State and Territory Statistics – 2019 ^35^ and were assumed to be 3 times more susceptible to infection with COVID-19 ^36^. The number of medical practitioners, nurses, midwives, and pharmacists in NSW is 135,379 – we assumed 80-90% wold be in the clinical workforce and require vaccination (125,000 vaccinations). As their average age is respectively 47, 44 and 41 years old ^35^, we distributed them through 8 age groups from 25 to 64 years old, following the age distribution of HCWs.

For ring vaccination we assumed varying proportions of vaccination of all the traced contacts - 70%, 80% and 90%. For use as PEP, we assumed a 50% reduction in efficacy (from 90% to 45%) when given to latent pre-symptomatic people.

For mass vaccination in NSW, assuming that the vaccine supply is available for the population, we tested the effect of speed of achieving high coverage by comparing the capacity to deliver 50,000, 75,000, 100,000, 125,000, and 300,000 doses per day to vaccinate the NSW population. We also varied the efficacy of the vaccine to determine the minimum vaccine efficacy to achieve herd immunity, and the epidemic scenario with low and high efficacy vaccines. We used the following formula to calculate the required vaccine coverage (Vc) for herd immunity assuming R0=2.5. ^28^ In the base case, vaccine efficacy (Ve) = 0.9 (90%).

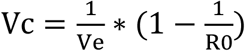

To test the effect of varied efficacy, vaccine coverage was held constant at 70%, reflecting a feasible mass vaccination goal. We tested varied vaccine efficacy of 38%, 50%, 60%, 70%, 80%, 90% and 95%. These estimates were selected based on published upper efficacy of vaccines of 94-95% ^9,10,12^, and the remaining values to reflect a plausible efficacy range of forthcoming vaccines. The lower estimate of 38% is the efficacy against any laboratory confirmed infection (symptomatic or asymptomatic) calculated from ChAdOx1 nCoV-19 vaccine (AZD1222) published data, excluding the data from the Brazilian component of the study, because weekly testing for infection was not done in Brazil. ^10^

Model outputs are the epidemic curve (cases), total number of cases and deaths and, for ring and mass vaccination, there is the added output of total number of vaccine doses used and days to vaccinate the entire susceptible population, respectively.

## Results

A restricted supply (for 1 million people) will not be enough to control the epidemic, with all scenarios resulting in a large number of cases and deaths after 500 days. Figure 1 shows results for the choice of using the available doses in the age group 10-29 years old, vaccinating HCWs and then 10-29 years old, or vaccinating the 65+ years old. Targeting the younger age group will have more impact on reducing the number of cases, whilst vaccinating people 65+ will have more impact on deaths, as this age group is at much higher risk of death.

**Figure 1:**
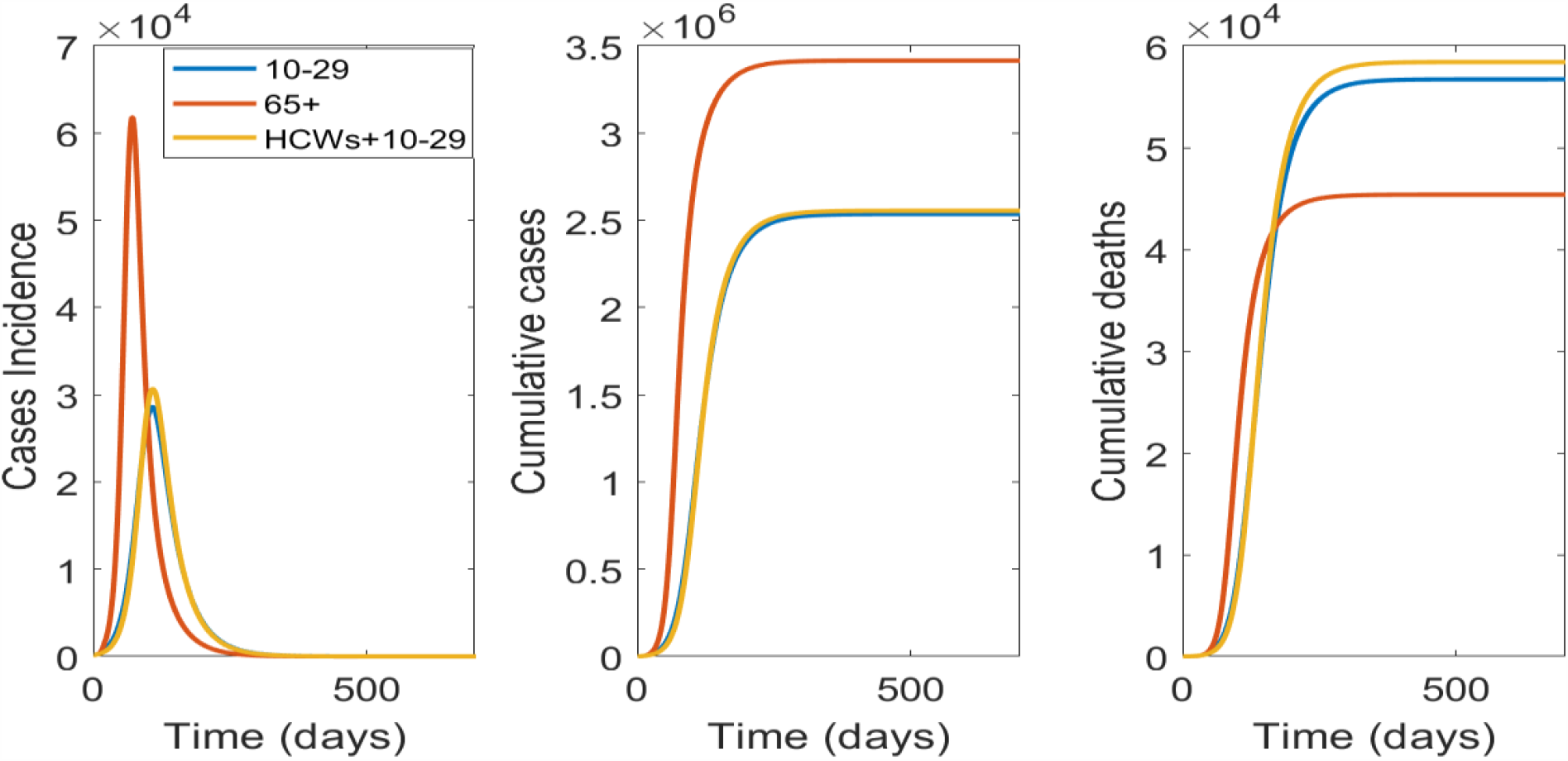
Targeted vaccination strategies with a restricted supply for 1 million people: From left to right is the epidemic curve, the cumulative case and deaths numbers by targeted age group vaccinated

If enough vaccine is available to vaccinate the entire population of NSW, the important variables influencing the results will vaccine efficacy and the speed of vaccination. In Figure 2 we show results of varying vaccination capacity from 50,000, to 300,000 people per day. Vaccinating 50,000 people per day will take about 177 days to vaccinate the entire population with one dose, will result in a much longer and larger epidemic, compared to 125,000 or 300,000 people per day. However, if the health system has capacity to vaccinate at least 75,000 people per day the epidemic could be terminated rapidly (Figure 2).

**Figure 2:**
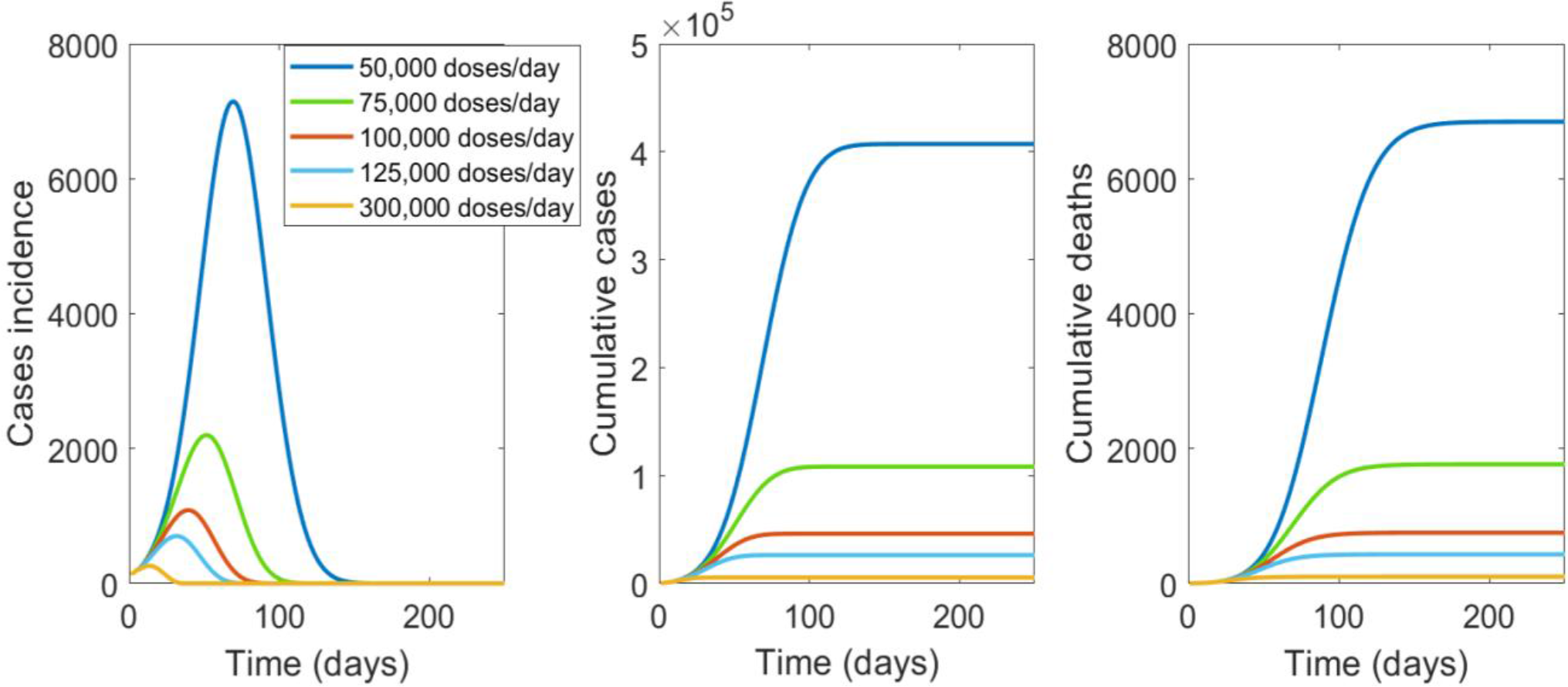
The impact of speed of mass vaccination with a vaccine of 90% efficacy. From left to right is the epidemic curve, the cumulative case and deaths numbers by number of number of people vaccinated per day in NSW.

With a vaccine efficacy (VE) of 90%, vaccinating 66% of the total population (approximately 5.384 million people) will provide herd immunity and reduce the R0 to less than 1. A minimum VE of 85% is required to achieve herd immunity at 70% population coverage. A VE of 80% will require 75% of the population to be vaccinated, and VE of 60% will require 100% of the population to be vaccinated to achieve herd immunity. Table 1 shows the required vaccine coverage for varying VE, and that efficacy below 60% cannot achieve herd immunity.

**Table 1:**
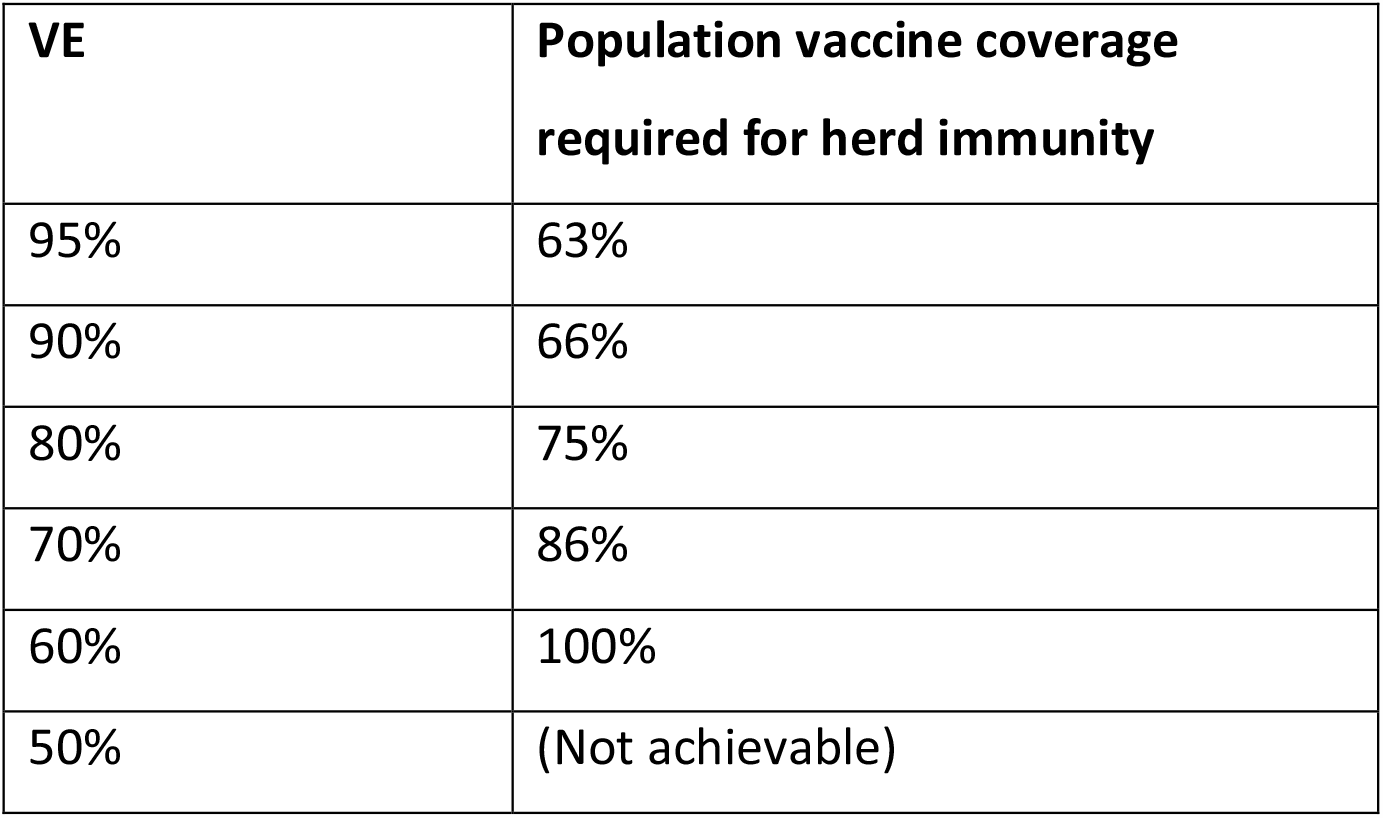
Vaccine coverage required for herd immunity by varying vaccine efficacy against all infection.

| VE | Population vaccine coverage required for herd immunity |
| --- | --- |
| 95% | 63% |
| 90% | 66% |
| 80% | 75% |
| 70% | 86% |
| 60% | 100% |
| 50% | (Not achievable) |

Figure 3 shows the effect of varied vaccine efficacy on the epidemic, with vaccine coverage of 70% (5.25 million people) with 125,000 doses delivered per day. When VE drops below 70%, epidemic control is substantially worse.

**Figure 3:**
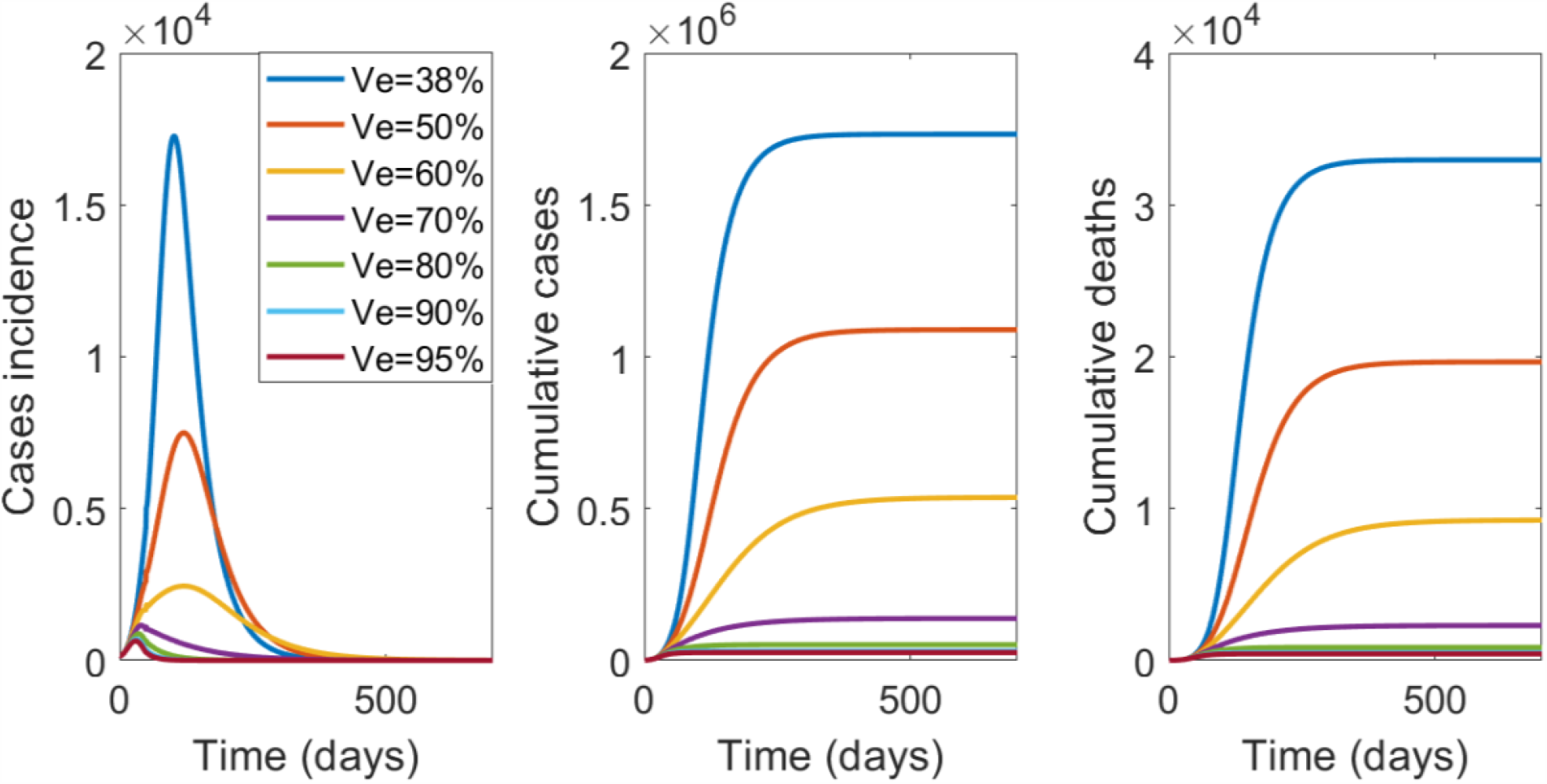
From left to right is the epidemic curve, the cumulative case and deaths by varying vaccine efficacy (VE), with 125,000 people per day vaccinated until 70% of NSW population is vaccinated.

In Figure 4 the results for ring vaccination with varying rates of contact tracing per day (70%, 80% or 90% of contacts of cases) are shown. The higher the proportion of contacts traced and vaccinated from the start, the lower the required vaccine doses. If only 70% of contacts are traced, the required vaccine doses are much higher and the epidemic impact much greater than 80 or 90% (Figure 3).

**Figure 4:**
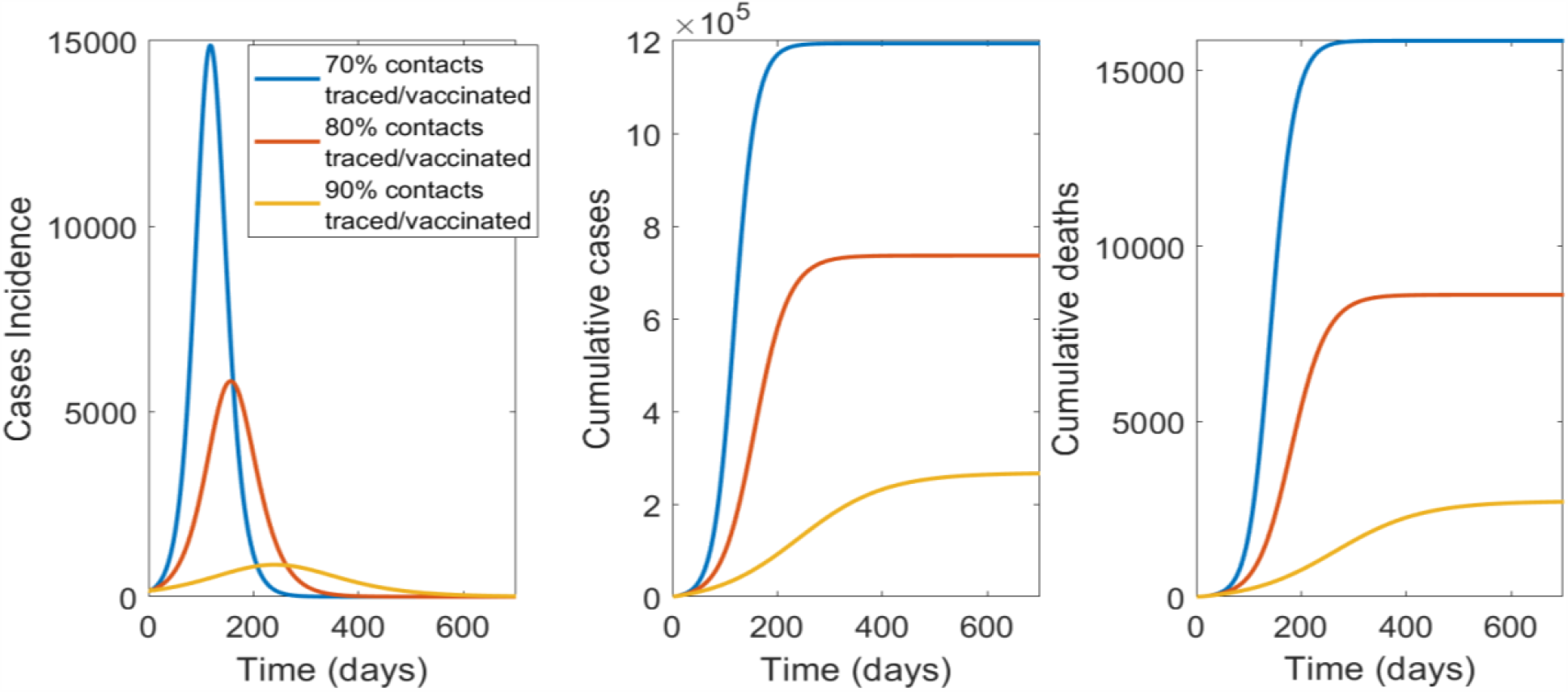
The impact of contact tracing and ring vaccination on epidemic control for a vaccine with 45% efficacy as post-exposure prophylaxis: From left to right is the epidemic curve, the cumulative case and deaths numbers by percentage of contacts traced and vaccinated

Table 2 shows all numerical results for the different scenario tested. In conclusion, with a limited vaccine stockpile the most efficient way to delivery vaccine doses will be ring vaccination, assuming a 45% efficacy when used as PEP, and at least 90% of contacts per case traced and vaccinated. If supply of vaccine is adequate to cover the entire NSW population, mass vaccination with a high efficacy vaccine and rapid uptake (at least 75,000 people vaccinated per day) will be the only strategy able to achieve herd immunity and prevent community transmission.

**Table 2:** Total cases and deaths for targeted and mass vaccination scenarios, and total cases, deaths and vaccination used for ring vaccination scenarios

| Targeted vaccination: age groups |  | Total cases | Total deaths |
| --- | --- | --- | --- |
| 1) 10-29 |  | 2,536,040 | 56,708 |
| 2) 65+ |  | 3,415,521 | 45,379 |
| 3)HCWs + 10-29 |  | 2,554,442 | 58,409 |
| Mass vaccination: people vaccinated per day (total 5,250,000 doses, 70% coverage) |  | Total cases | Total deaths |
| 50,000 |  | 407,198 | 6,845 |
| 75,000 |  | 108,233 | 1,766 |
| 100,000 |  | 46,073 | 752 |
| 125,000 |  | 26,197 | 431 |
| 300,000 |  | 5,507 | 100 |
| Contacts traced/vaccinated | Total doses used | Total cases | Total deaths |
| 70% | 2,494,500 | 1,194,140 | 15,850 |
| 80% | 1,849,300 | 736,254 | 8,608 |
| 90% | 794,720 | 266,164 | 2,707 |

## Discussion

We showed that herd immunity through vaccination is the only means to preventing sustained community transmission and requires mass vaccination of 66% of the population with a vaccine of 90% efficacy. Vaccines of lower efficacy require higher vaccination coverage – 100% if the VE is only 60%. Vaccines with less than 60% efficacy cannot achieve herd immunity. Mass vaccination with a high efficacy vaccine represents the best strategy for economic recovery, as herd immunity through effective vaccination will provide the best prospect for lifting of societal restrictions. Elimination cannot be achieved and sustained without mass vaccination ^37^.

We show that for a population of 7.5 million people, if faced with an initial restriction in vaccine supply, sustained epidemic control cannot be achieved by vaccination alone, and other non-pharmaceutical interventions will need to continue to mitigate the risk of outbreaks ^2^.If COVID-19 vaccines have efficacy as PEP, contact tracing and ring vaccination is the best way to achieve epidemic control, but the majority of the population will remain non-immune and susceptible to outbreaks. Recurrent breaches in hotel quarantine ^38^ and associated with cruise ships have resulted in community transmission, and without herd immunity, this risk will continue and will hamper economic recovery ^39^.

During the period of initial restricted supply, health and aged care workers should be the highest priority, as protecting health workers and other first responders is essential for health system resilience in a pandemic. During a large second wave in the state of Victoria, over 3500 health and aged care workers became infected ^40^, and we demonstrated ongoing risk to health workers, even during periods of low community incidence ^36^. After vaccinating health and aged care workers, the evidence supports vaccination of older adults. Age is the strongest predictor of morbidity and mortality, far more than any chronic disease ^41^, so protecting older adults will prevent the most deaths from COVID-19. Including younger people in the prioritisation group can reduce transmission and the overall infection rates, although this benefit is negated by the higher mortality rates in older people. Those findings are supported from a preprint study, which explored the most effective age-group to prioritize for vaccination in order to reduce morbidity and mortality from Covid-19 ^42^.

If COVID-19 vaccines are effective as PEP, this will be the best approach for epidemic control using a limited vaccine supply and requires capacity for high levels of contact tracing. If community transmission of COVID-19 remains low, complete contact tracing and ring vaccination will be a highly cost-effective, dose-sparing strategy. Whilst there are no data yet on the efficacy of COVID-19 vaccines as PEP, we showed that if a 50% reduction of efficacy is seen with these vaccines, that ring vaccination can be an effective strategy, and may be a better use of a restricted supply initially, if rates of contact tracing are high. Many vaccines have efficacy as post-exposure prophylaxis in outbreak settings, including measles, hepatitis A, varicella zoster and smallpox, with an efficacy of 50-90% of that for primary prevention ^20^. This is a research gap for COVID-19 vaccines, which needs to be addressed. If a high (>80%) percentage of contacts cannot be traced (such as when an epidemic is very large and the scale of contact tracing is unmanageable, as occurred in the second wave in the State of Victoria ^43^, mass vaccination becomes more urgent.

We showed that for mass vaccination, a slow trickle approach will leave us living with COVID-19 longer with all the associated societal and economic implications - rapid speed of vaccination will be the best strategy to reduce morbidity and mortality. The logistics of rapid delivery is key, and we show that slower uptake will result in more cases and deaths and a longer epidemic risk. Rapid vaccine uptake requires concerted planning for vaccination infrastructure, including training more accredited vaccinators, planning mass vaccination clinics, and testing the capacity to deliver 300,000 or more vaccine doses a day, which provides the best outcomes. We assumed in the base case scenario for mass vaccination that the health system will be able to deliver 125,000 doses per day, which is similar to the distribution capacity estimated in Victoria of about 117,000 doses per day ^44^. We showed that the speed of Covid-19 vaccine uptake is critical to rapid epidemic control, however the logistics of vaccine distribution is complex ^44^, particularly for a country such Australia, where an estimated 29% of the population lives in rural and remote areas ^45^. Therefore, development of detailed logistical plans and tools to support increased capacity to vaccinate, as well as effective vaccine transport, storage, and continuous cold-chain monitoring is required.

No country has attempted mass vaccination since the eradication of smallpox. Currently in Australia, vaccination is delivered mainly in primary care. Under the National Immunisation Program about 5 million doses are delivered across the country each year in the infant, adolescent and adult vaccination programs. Mass vaccination requires this to be scaled up by five times. Nurse vaccinators can add to the primary care capacity of general practitioners but require accredited training to be able to vaccinate. In addition, pharmacists can vaccinate for influenza in Australia and may be an additional source of accredited vaccinators, but giving a new vaccine with uncertain safety profile for rare adverse events would be best done in general practice or in public vaccination clinics staffed by medical and nursing personnel. Modelling of GP capacity to rapidly vaccinate 70-80% of the population, and the impact on other medical care, is needed, and logistic support and resourcing for GPS to expand their capacity to achieve this will be required. This includes cold chain infrastructure for vaccines that require deep freezing ^12^. Decisions need to be made on public vaccination clinics, staffing, training of more nurse vaccinators and other measures to increase vaccination capacity.

Australia does not have a goal of herd immunity through mass vaccination at this stage, which means uptake will not likely be adequate for herd immunity. A vaccine with 90% VE requires 66% coverage, which is feasible, even without children being vaccinated. At this stage, because of lack of phase three clinical trial data in children, initial vaccine roll out will be restricted to adults, who comprise 80% of the population. This also makes age-based targeted vaccination only feasible for older adults. However, adult vaccination is more challenging than infant vaccination, as adults are a mobile population – for people over 65 years, about 70% receive influenza vaccine annually. ^46^ Achieving whole-of-life vaccination rates above 70% may be challenging, and impossible while COVID-19 vaccines remain unlicensed in children, who comprise 20% of the population. This is another impetus for choosing a vaccine with at least 90% VE, as the required coverage for herd immunity (66%) is far more feasible, even if children are not vaccinated.

Vaccine hesitancy may also affect the ability to achieve rapid, high uptake and must also be addressed well in advance of vaccination programs to ensure that programmatic competency to achieve high uptake rapidly will not be met by unexpected social or behavioral obstacles. Australia has very high rates of vaccination for National Immunization Program vaccines ^47^, especially with the use of financial disincentives, and has a more accepting culture of public health interventions, so large scale vaccine refusal is not expected. We also identified that vaccine hesitancy about COVID-19 vaccines mostly does not reflect fixed objection to vaccination, and that there is scope and need for health promotion and positive messaging to ensure high vaccine uptake ^48^.

In conclusion, restricted vaccine supply can protect first responders but will not have an impact on epidemic control, unless the vaccine has some efficacy as PEP. If vaccines are effective as PEP, then vaccinating health workers and ring vaccination would be the preferred use of a restricted supply. We recommend that clinical trials urgently be conducted to study the efficacy of COVID-19 vaccines as PEP given to close contacts of cases. When vaccine supply is available for mass vaccination, the quicker a high uptake can be achieved, the greater the impact on epidemic control, morbidity and mortality, but herd immunity cannot be achieved with vaccines of less than 60 % efficacy. The VE of the ChAdOx1 nCoV-19 vaccine, the largest component of the Australian planned stockpile, is less than 40% against any laboratory confirmed infection (symptomatic or asymptomatic) based on interim analysis. ^10^ The use of high efficacy vaccines provides the only prospect of achieving herd immunity against SARS-COV-2, and such vaccines should be procured if herd immunity is a goal. There are no data on serial dosing with two different vaccines, which is a possible scenario if Australia starts with a lower efficacy vaccine. This is another reason to ensure adequate supply of high efficacy vaccines. The logistics of mass vaccination, including cold chain, delivery, capacity to vaccinate and addressing vaccine hesitancy should be a high priority for planning and resourcing.

## Data Availability

All data are from published studies, available in the reference list.

## Supplementary information

### Model Description

We used three different model structures to simulate targeted, mass and ring vaccination strategies. Matlab 2019 software was used to build deterministic compartmental disease transmission models. We used an expanded SEIR system of ordinary differential equations, where the population (the initial susceptible) is divided in 16 age groups, 5 years each from 0 years old to 74 years old and an additional age group comprising people 75+ years old. Furthermore, the population is divided in two groups, health care workers (HCWs) and general population, to take into account the higher infection risk for HCWs. The differential equations move the population through diseases epidemiological stages, as susceptible (S), latent not infectious yet (E) latent infectious undiagnosed (E^u^), contacts traced and isolated for 14 days (C), symptomatic infectious stages for undiagnosed (I1,I11) and diagnosed (I2,I22), recovered (R) or death (D), and public health response stages, as latent infectious diagnosed (E^t^) or isolated (Q), cases hospitalized (H) and requiring intensive care unit (ICU), and the vaccinated compartment (V). The H and ICU outputs are not shown in the outputs, but are assumed to have reduced transmission risk in the model once cases are isolated. We have two compartments for asymptomatic people, (A1 and A2). A1 represent the peak of infectiousness and A2 the 6 following days of gradually decreasing infectivity.

The force of infection, which is the age specific rates at which susceptible people get infected is described as

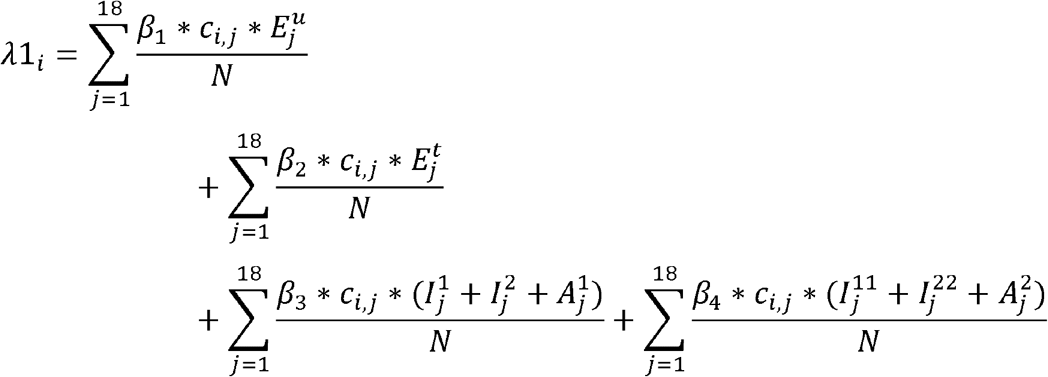

Where 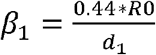 for latent undiagnosed contacts, 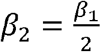 for latent diagnosed and home quarantined (50% reduction in R0), *β*_3_ = 0.36\**R*0 for the first day of symptoms an 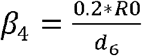 for the following 6 days of symptoms. *c*_*i,j*_ is the age-specific contact matrix adapted from (1) for New South Wales, and *N* is the total population.

While the rate of age specific contacts that are not infected are calculated as

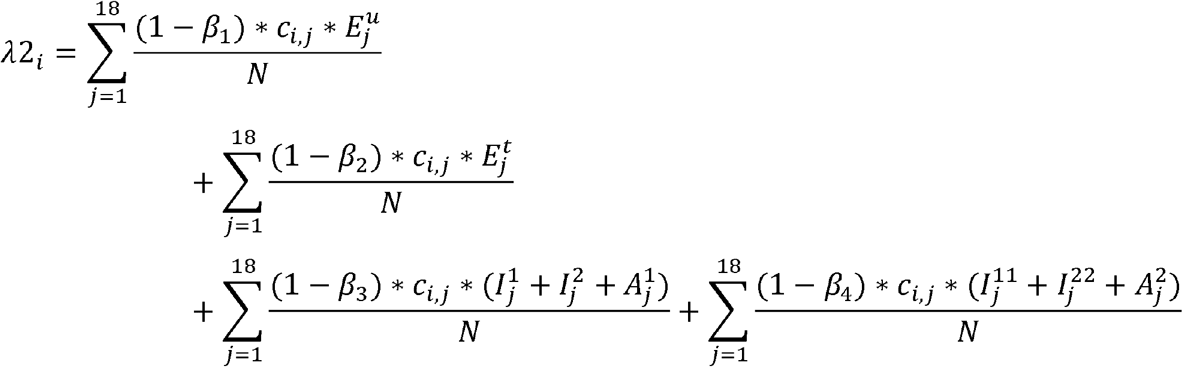

### Targeted and mass vaccination models

For the implementation of targeted and mass vaccination strategy for outbreak response we used the same model structure, however we used different matrix X_j,i_ for vaccination rates between each targeted strategy and mass vaccination. In the scenario of age-targeted vaccination, we included only susceptible people and the vaccine distribution rates are constantly distributed equally every day to the same targeted age group until the available stockpile is finished. For mass vaccination the distribution rates in age groups and epidemiological state are directly proportional to the number of susceptible and latent people each day.

If M_j,i_ is the matrix by 2 epidemiological states (S and E) and by 16 age groups, containing all the population susceptible and latent who will get vaccinated, and X_j,i_ is the same size matrix of vaccine distribution rates, each element *x*_*j,i*_, of the matrix X_j,i_ is recalculated each day as an endogenously estimated parameter, which depends on the number of people in each of the compartments *m*_*j,i*_, of the matrix M_j,i_. In formula each element *x*_*j,i*_, of the distribution matrix X_j,i_ is recalculated every day as

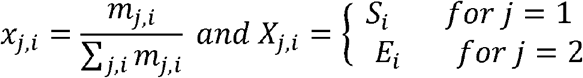

*m*_*j,i*_ is the number of people in each compartment of the matrix M. All contacts are assumed to be quarantining for 14 days.

Following are the differential equations used for these two models with *i=1,…,16* age groups,

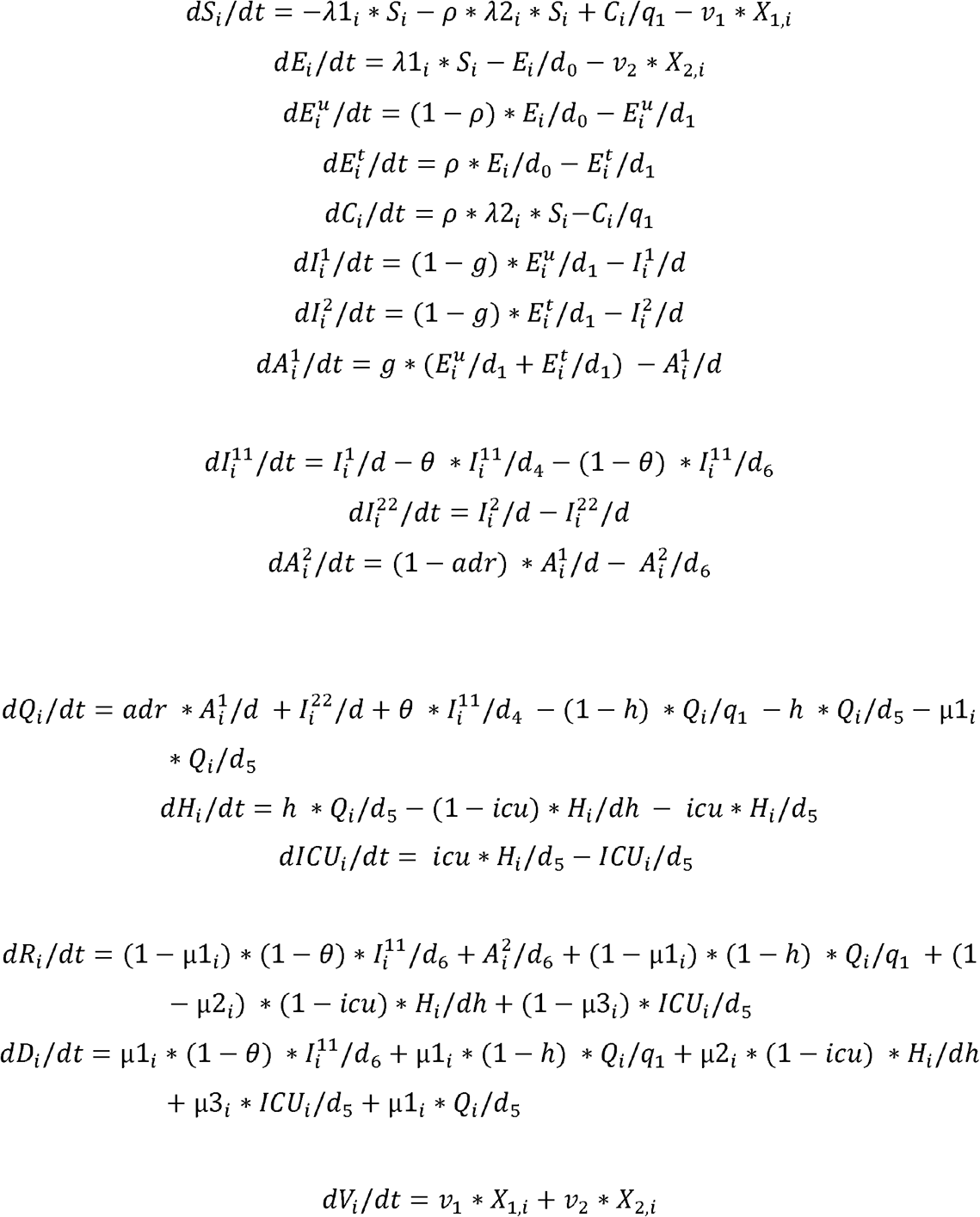

### Ring vaccination model

In the ring vaccination scenario, the contacts traced are vaccinated in addition to being quarantined. The following are the differential equations used

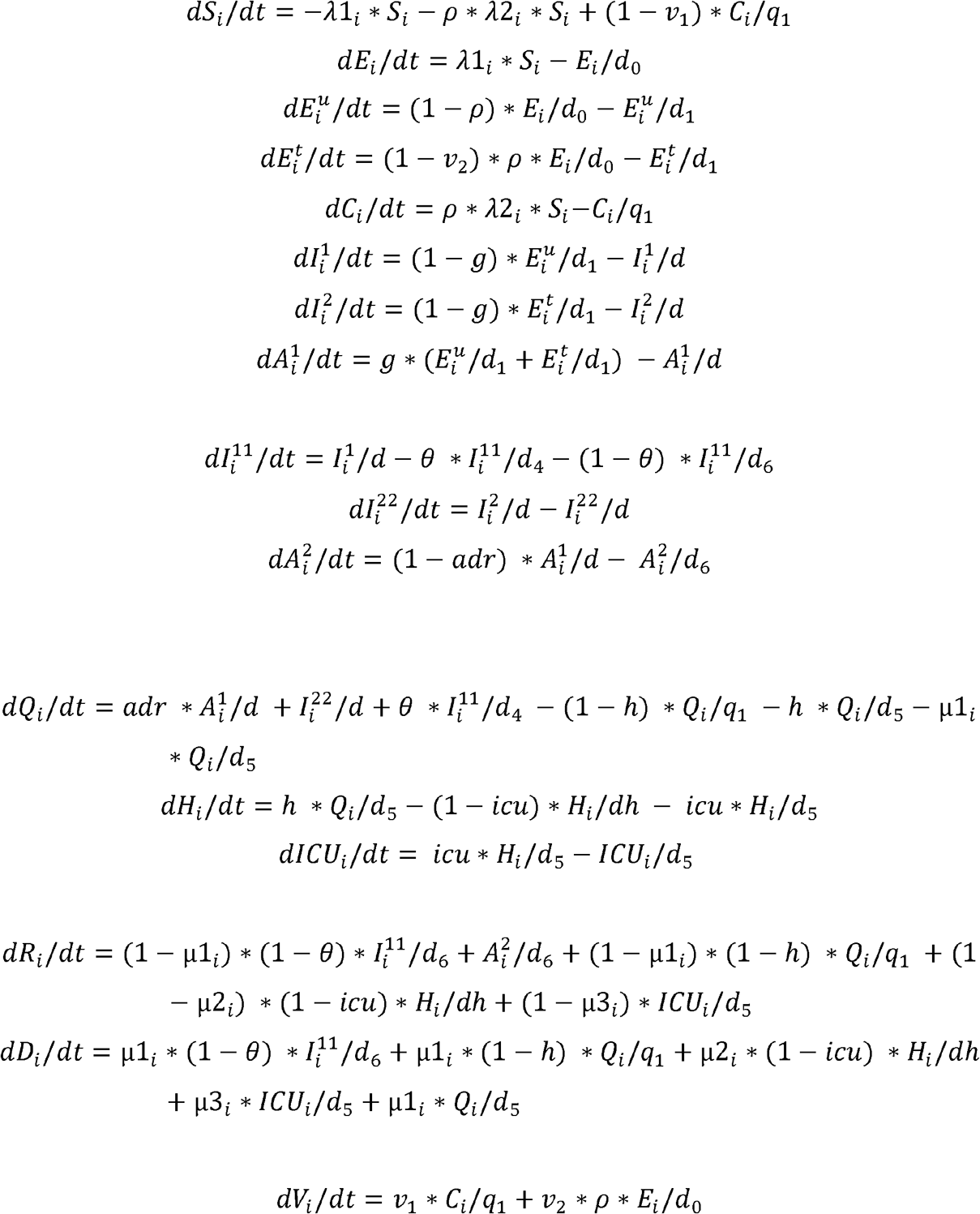

### Parameters table

**Table S1:**
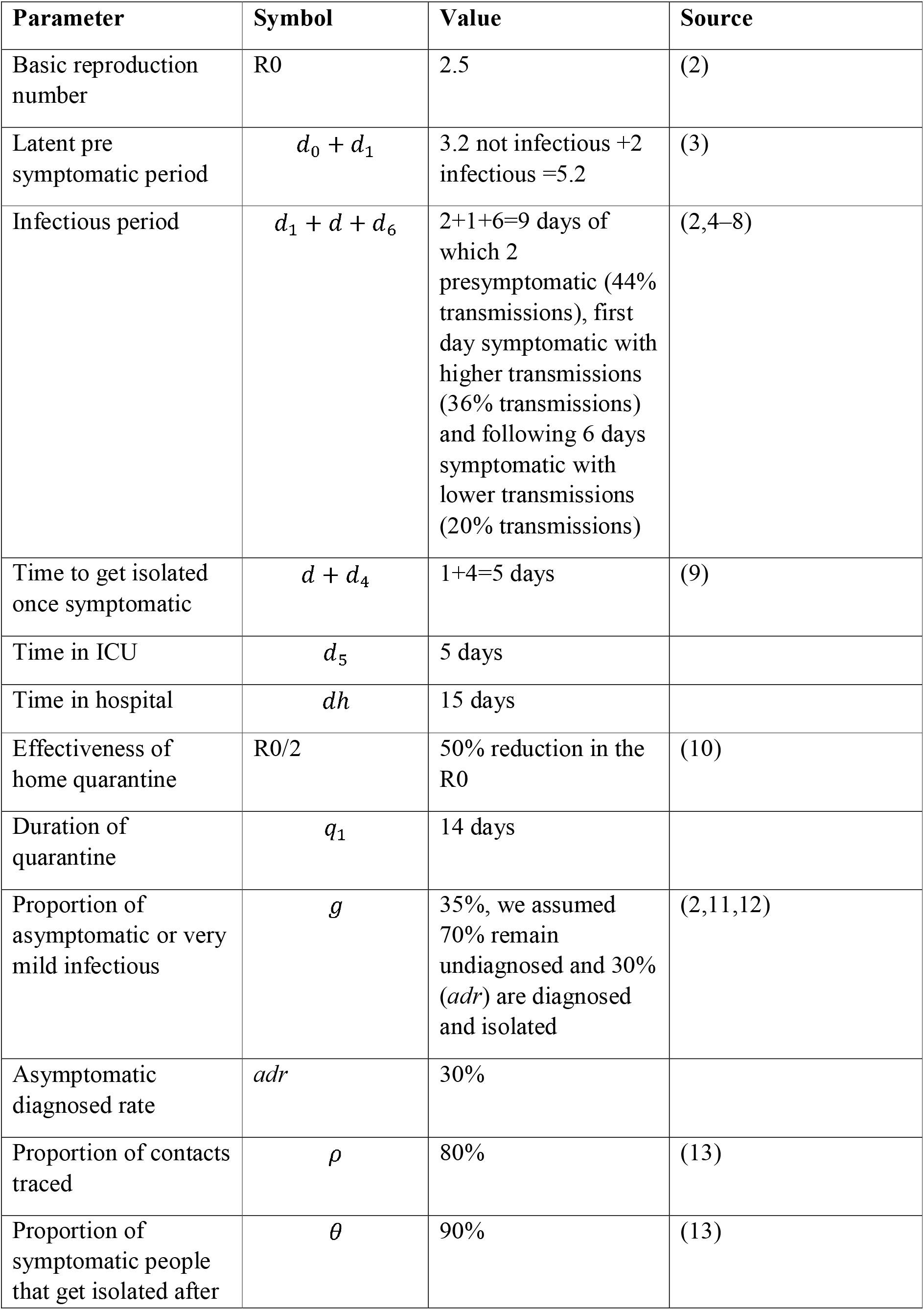
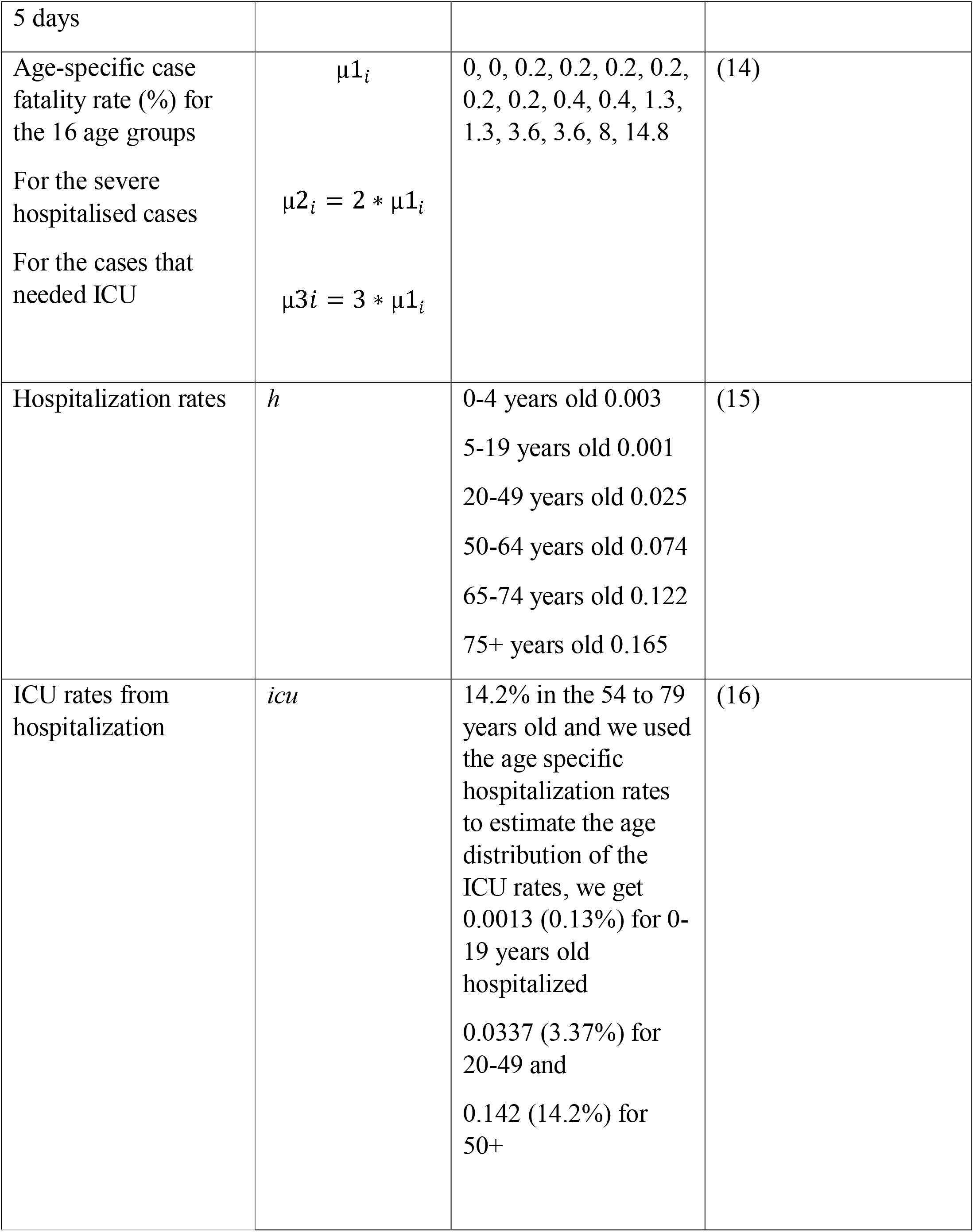
model parameters used and their values

